# Checking the calculation model for the coronavirus epidemic in Berlin. The first steps towards predicting the spread of the epidemic

**DOI:** 10.1101/2020.11.14.20231837

**Authors:** D. Below, J. Mairanowski, F. Mairanowski

## Abstract

A calculation model has been proposed to forecast the spread of the СOVID-19 epidemic under quarantine conditions. The resulting simple analytical relationships allow for the assessment of factors determining the intensity of the spread of infection, including the changing requirements for quarantine severity. The prediction method presented makes it possible to calculate both the total number of infected persons and the maximum rate of spread of infection.

Following the publication of this work in May 2020, in October this year there was a new surge in the virus epidemic, the intensity of which depends on the population’s compliance with the rules of hygiene and social distance. Comparison of the results of the model calculations with the statistics for Berlin shows that they are of satisfactory quality. In particular, it shows that with an epidemic growth rate of around 1,000 people/day, unless additional quarantine measures are taken, the total number of infections can be expected to approach 100,000 within approximately six months. It is shown that the intensity of the virus’s spread depends on the socio-demographic composition of the population in different districts of Berlin and age structure. The possible impact of behavioural factors dependent on the psychological state of people on the spread of the epidemic, which can be assessed by analysing changes in heart rate, is discussed.

---

Since the release of our work [1] in May 2020, i.e. during the initial period of the spread of COVID 19, the dynamics of the epidemic have, as might be expected, been largely determined by the degree of rigour in implementing protective measures such as limiting contact, wearing masks in transport and other public places, maintaining social distance, etc.

As a result of the reduced intensity of the spread of the epidemic, a number of restrictions on the operation of businesses, schools, kindergartens and various institutions in Berlin have been lifted. During the summer, large numbers of Berliners went on holiday to countries at high risk of infection, such as Turkey, Spain, Italy and France.

In mid-August, Berlin schools and institutions of higher education started working on schedule again. All of this led to an increase in the rate of infection.

In work [1], correlations were obtained to calculate the total number of infected coronavirus patients, including in the context of softening quarantine requirements. Calculations were made on these ratios, which showed, among other things, that depending on the level of quarantine mitigation and the relative number of people with congenital or acquired immunity to the disease, the total number of diseases in Berlin could reach 200,000 or even 400,000. Figure 1 shows the graph resulting from this calculation [1] for the conditions in which the measures to limit the spread of the epidemic will be practically eliminated.

**Figure 1:**
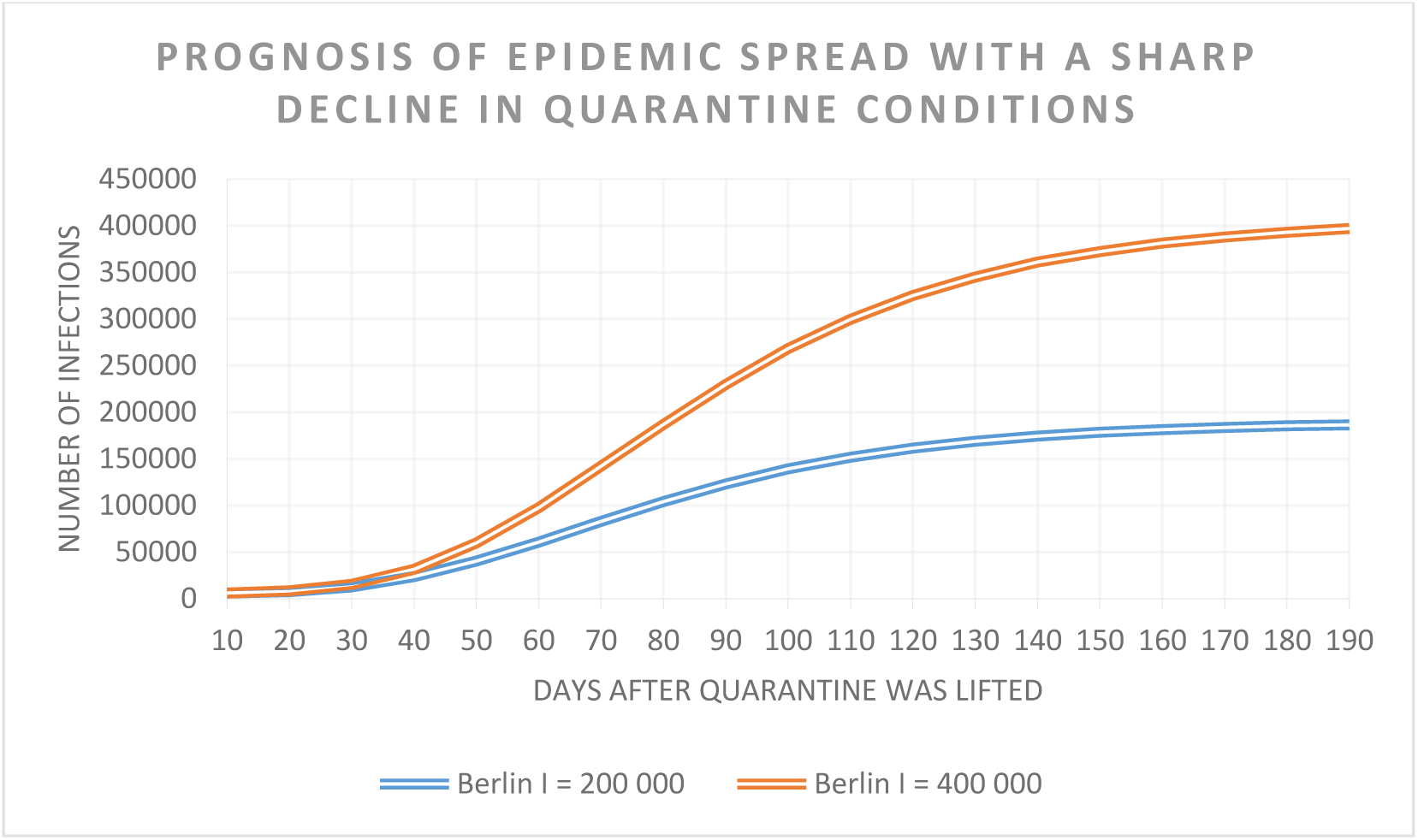
Calculation of the total number of diseases according to [1].

In addition to calculating the total number of people becoming ill, it was also proposed to calculate the time of the maximum daily increase in the number of infected people, counting from the beginning of quarantine weakening:

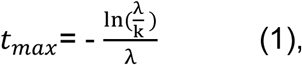

in which:

λ is a factor of decrease in contact with infected patients with persons who may potentially get infected by means of quarantine and other preventive measures,

k is a factor that characterises the speed of transmission of the virus, depending on both the nature of the virus and the infrastructure in the population centres and the characteristics of the social and demographic situation. For Berlin, according to [1], this factor was taken as k = 0.4 1/day.

This continuation of [1] looks at the characteristics of the spread of the virus epidemic in a context of partial quarantine weakening, using the example of Berlin from 14^th^ of August 2020. Statistical data on the number of infected patients between mid-August and mid-November are used to check the results of the calculations.

The main correlation used to predict the growth of the epidemic by type coincides with the ratio (4) on which the epidemiological situation in Berlin was assessed. It will be written down in the following form:

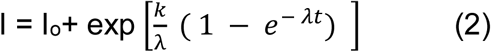

Io - number of infected patients at the beginning of the settlement period,

t - time in days counted from the beginning of the settlement period.

The difference between the ratio (2) and a similar equation presented in [1] is the assumption that a new epidemic is mainly associated with a modified strain of the virus. This assumption is based on the work of virologists who have established that, since midsummer, the epidemic in Europe has been associated mainly with a new strain of the 20A.EU1 virus [2].

Analysis of data on the number of actively infected persons shows that, for the Berlin context, the number of cured patients depends little on the total number of infected people, amounting at the beginning of the epidemic in question to around 200 people per day, and subsequently increasing to between 500 and 600 people per day. It is therefore easy to calculate the number of actively infected patients, taking into account the real intensity of treatment, which depends on the level of the city’s health care system.

The start of the calculation period is based on the date, the active return of a large number of holiday-makers from holiday and the start of classes in schools. This date was taken into account on 14^th^ of August. Accordingly, the calculation was based on the actual data for Berlin Io = 10,000. The calculations were made without taking into account the treatment of patients with the virus.

The time of the maximum intensity of growth of patients was determined by (1). By substituting (1) for (2), we obtain a ratio for maximum epidemic growth (1/day):

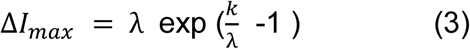

As before, the calculation was based on a coefficient k = 0.4 1/day.

As for the coefficient λ, its value depends on the strict observance of social distance and the restrictions on people-to-people contacts accepted. Depending on the choice of this coefficient, both the total number of infected patients changes, *I*_*max*_, and the maximum rate of spread of infection, Δ*I*_*max*_.

Table 1 presents the results of the calculations of these values in terms of ratios (2) and (3) respectively.

**Table 1:** Results of the calculations of the values Δ*I*_*max*_, *I*_*max*_ and *t*_*max*_ in terms of ratios (2) and (3).

| $\lambda$ | 0,040 | 0,039 | 0,038 | 0,037 | 0,036 | 0,035 |
| --- | --- | --- | --- | --- | --- | --- |
| $\Delta I_{max}$ | 320 | 410 | 520 | 675 | 890 | 1200 |
| $I_{max}$ | 32000 | 38000 | 47000 | 60000 | 80000 | 100000 |
| $t_{max}$ | 58 | 60 | 62 | 64 | 67 | 70 |

The analysis of this table shows, in particular, that the maximum growth rate of infected patients observed at the beginning of November (more than 1000 people per day) corresponds to the value of the coefficient λ=0.035 - 0.036.

With these values, the maximum number of infected people in Berlin, according to Table 1, can reach between 80,000 and 100,000 people. Of course, provided that restrictive measures do not stabilise the spread of the epidemic.

Figure 2 shows the results of calculations from (2) for various contact restriction options and statistical data on the total number of infected patients in Berlin.

**Figure 2:**
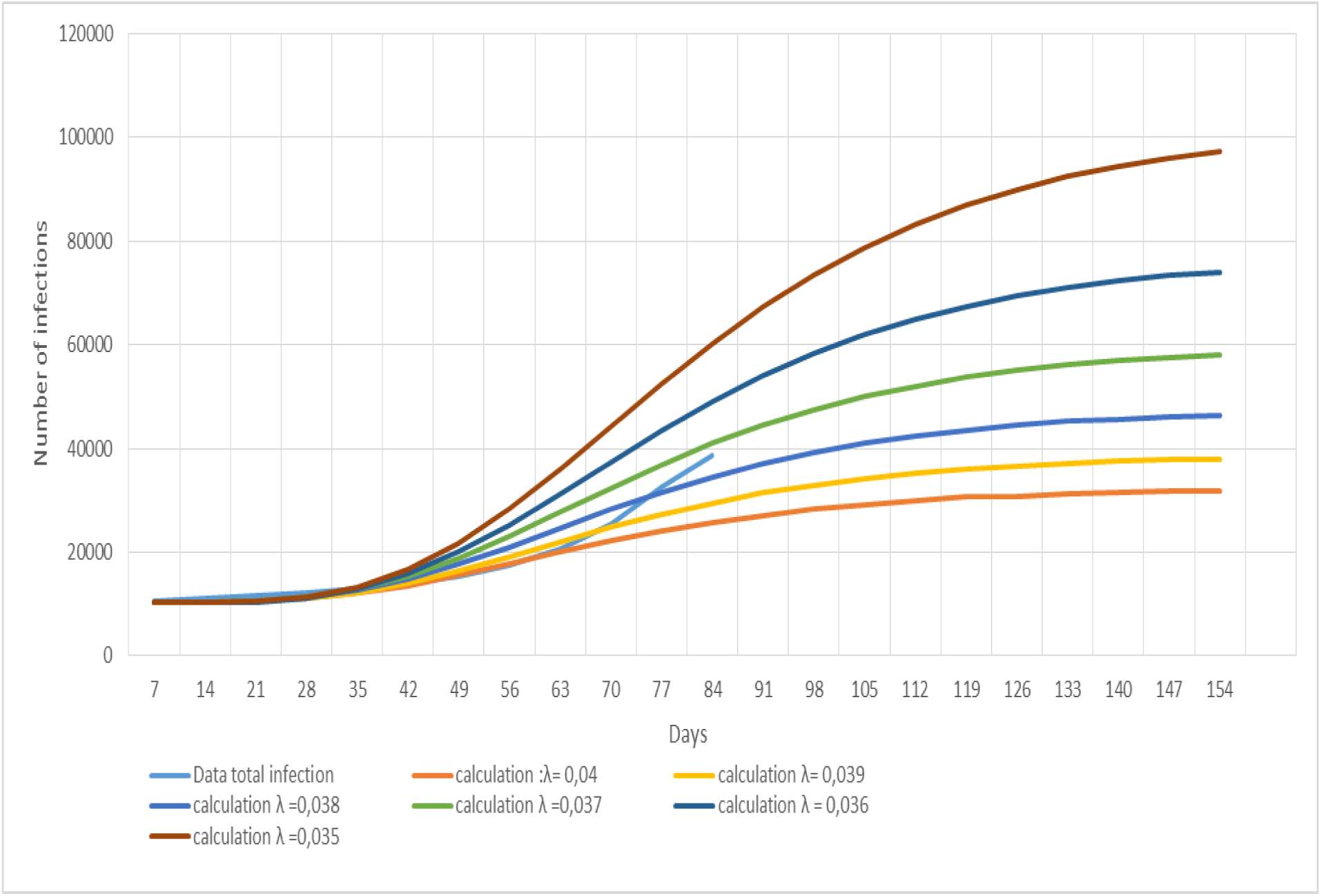
The spread of the coronavirus epidemic cumulative infection after partial lifting of quarantine restrictions.

The spread of the coronavirus epidemic cumulative infection following the partial lifting of quarantine can be seen in this figure, which shows that around two months after the quarantine was lifted, the epidemiological situation in Berlin has sharply deteriorated. This is consistent with the calculated time t_max_ at which the maximum growth rate of infected persons is predicted according to the addiction (3). At the time of writing, the maximum number of newly infected persons reaches 1200 per day. If this scenario were to develop further, the total number of infected persons would be expected to reach 100,000 by the end of the epidemic in Berlin. In this scenario, the epidemic will almost end only after 200 days, when the daily increase in the number of infected people will not exceed 15. If control of the epidemiological situation is lost, the scenario described in the first part of this paper may be possible, where the maximum increase in the number of sick people per day reaches 2000 and the total number of infected people reaches 200000 (see Fig. 1).

However, stricter quarantine measures introduced since November may cause a decrease in the growth rate of infected people and return to one of the more acceptable scenarios presented in Figure 2.

Thus, the results of the forecast depend on the choice of an epidemic scenario and this does not allow for a scientifically substantiated unambiguous forecast regardless of the accuracy and rigour of the task definition in the model development.

The further development of the model should thus go in the direction of identifying causal links between the intensity of the epidemic and the main factors affecting this process. The absence of such a link prevents the development of reliable and unambiguous forecast models and forces the calculation to assume different epidemic scenarios. At the same time, the information already available to date makes it possible to take the first steps towards a scientifically sound choice of a scenario for the forecast model. Some of these factors are related to the characteristics of the population’s behaviour and the infrastructure of cities, while others are related to the characteristics of the virus’s spread, the development of natural or artificial immunity to the virus, and the effectiveness of treatment of the disease.

Furthermore, the impact of socio-demographic characteristics of the city’s population on the rate at which infection spreads should be considered.

A detailed analysis of the spread of the epidemic in Berlin shows that the number of infected people in the city is very uneven. For example, in some districts, such as Treptow-Köpenick, the maximum number of infected people does not exceed 50 per day, while in Neukölln it reaches 300 people per day or more. Fig. 3 presents statistical data on the growth rate of the epidemic in Berlin and the three characteristic districts of the city. On the horizontal axis, the days since the start of the new epidemic have been postponed; on the vertical axis, the ratio of infected residents in each district to the total population of the respective district as a percentage. The same graph shows the data for Berlin as a whole. The intensity of the epidemic in the Neukölln district was the highest not only in Berlin but also in other regions of Germany. In addition to the higher population density, these high figures in the area are explained by some of the characteristics of the behaviour of the inhabitants of the area, the large number of migrants from Turkey and Arab countries. Thus, outbreaks of the viral disease have often been recorded after the traditional crowded celebrations of various events. At the same time, it was not possible to identify the chain of participants in these celebrations, which prevented further mass infection.

**Figure 3:**
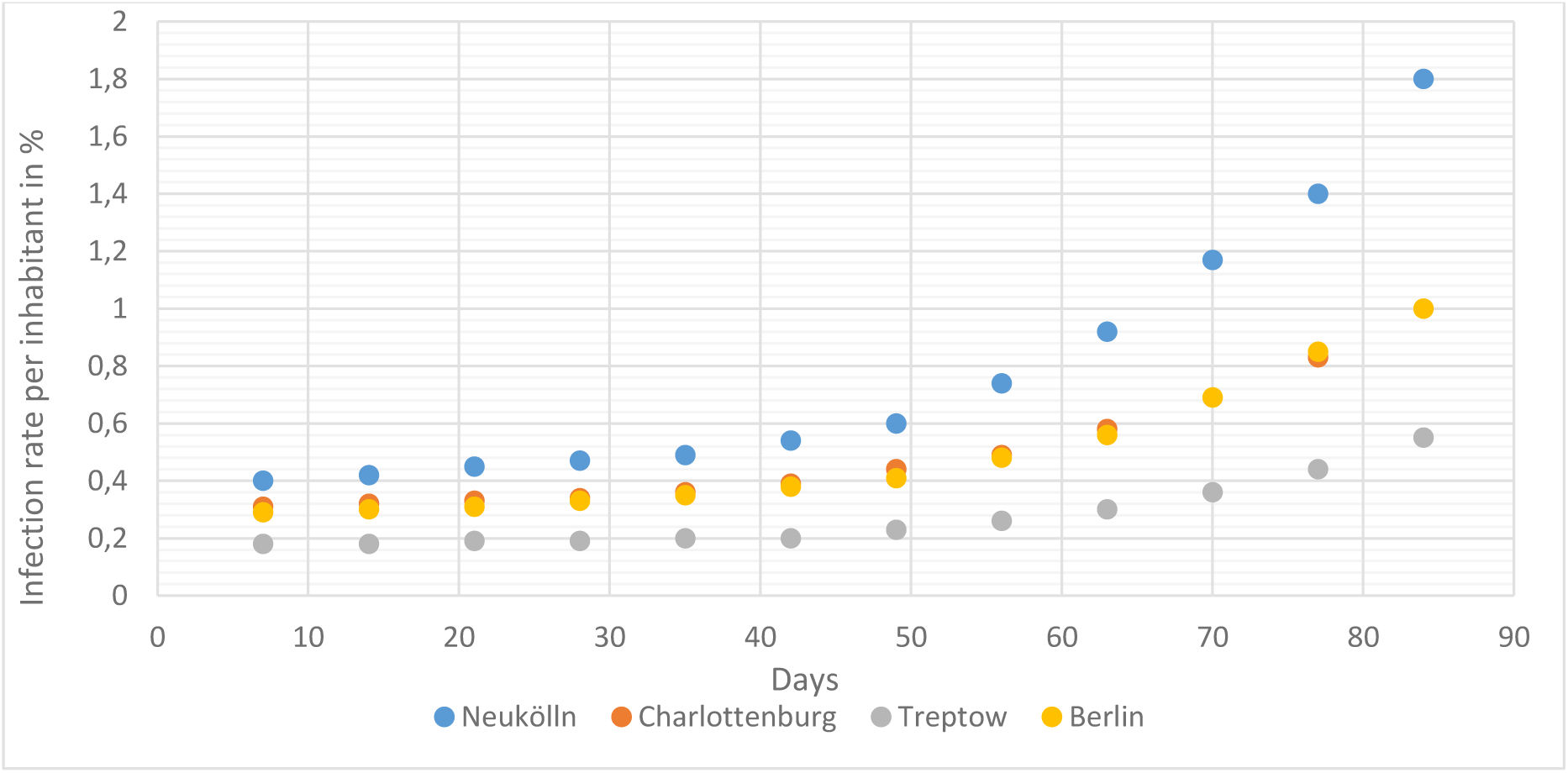
Intensity of epidemic development for selected districts of Berlin.

It is interesting to note that while for Neukölln the relative number of residents from Islamic countries is more than 20%, for the Charlottenburg-Wilmersdorf area it does not exceed 12%, which is roughly the same as the average for Berlin. For the Treptow-Köpenick area it is less than 5%. The data for Berlin and the district of Charlottenburg-Wilmersdorf are almost the same, while for Treptow-Köpenick the intensity of the epidemic is approximately 4 times less than for Neukölln and 2 times less than for the whole of Berlin.

These high figures in the Neukölln area can be explained not only by the higher population density, but also by some of the characteristics of the behaviour of the inhabitants of the area, the large number of migrants from Turkey and Arab countries. Thus, outbreaks of the viral disease have often been recorded after the traditional crowded celebrations of various events. At the same time, it was not possible to identify the chain of participants in these celebrations, which prevented further mass infection.

This can be explained by outbreaks of the viral disease which have often been recorded after the traditional crowded celebrations of various events. It was not possible to identify the chain of participants in these celebrations, which prevented further mass infection. In addition to the socio-demographic characteristics of the city’s population, the psychological state of people may also influence the spread of the epidemic.

Figure 4 shows the dependence of the number of infected people for each of the 12 districts in Berlin on the relative size of the population from the member countries of the Organization of the Islamic Conference (OIC) (Turkey, Arab States, some African countries). Statistics on the number of infections were taken for 1 November 2020 according to [3], data on the demographic composition of the inhabitants of each district according to [4]. The high correlation coefficient (r = 0.93) shows that the connection between these parameters is not only statistically reliable, but also practically functional. For the three districts with high percentages of foreign residents of these countries (Neukölln, Friedrichshain–Kreuzberg and Mitte) the number of infected persons exceeds 3 times the number of districts such as Treptow-Köpennik or Marzahn–Hellersdorf.

**Figure 4:**
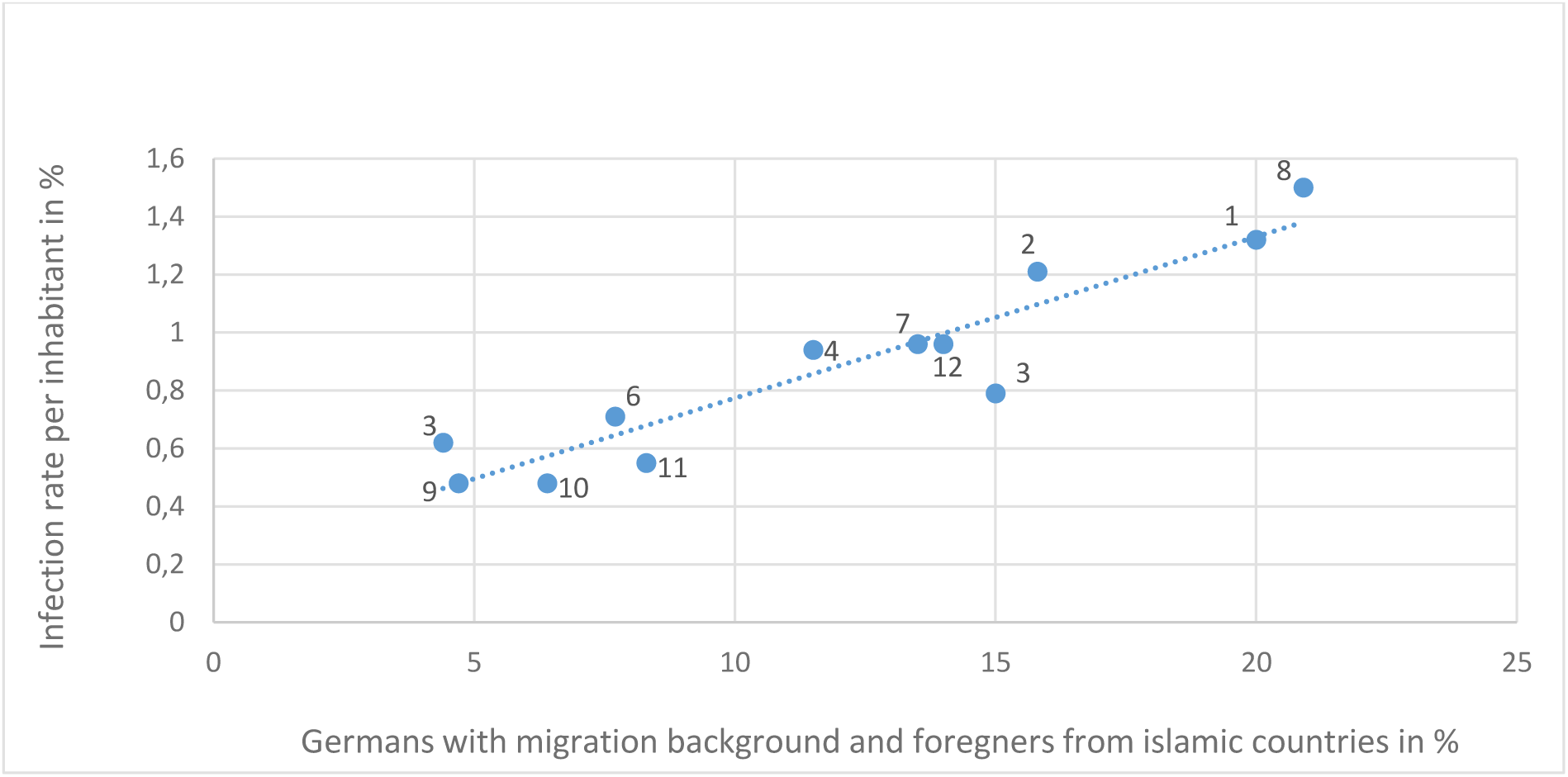
The dependence of infection rate on foreigners from countries OIC.

Figure 5 shows a graph of the correlation between the number of infected people in the same areas of Berlin and the number of foreigners from non-Islamic countries (EU countries, former Soviet Union, Vietnam, USA, etc.). There is also some dependence, but it is much weaker. (Correlation coefficient r = 0.66).

**Figure 5:**
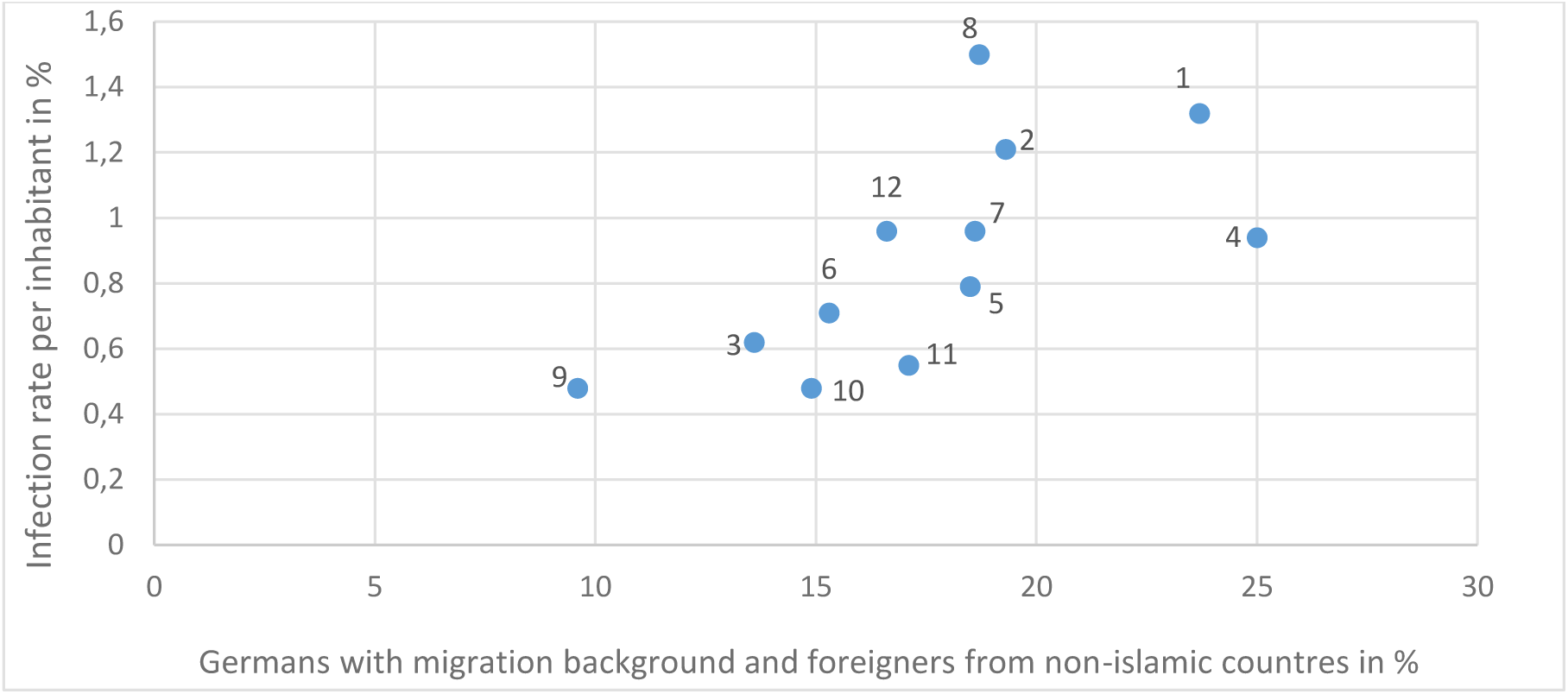
The dependence of infection rate on foreigners from non- Islamic countries.

In areas with high rates of infected patients, quarantine disorders, including violation of social distancing, are the highest in Berlin. These violations are so widespread that it has become necessary to use Bundeswehr soldiers to patrol the streets in the Mitte and Friedrichshain-Kreuzberg districts.

It is natural to assume that the intensity of distribution is influenced by population density, which varies considerably from one area to another. Official statistics [5] were also used to construct this dependency graph in Figure 6.

**Figure 6:**
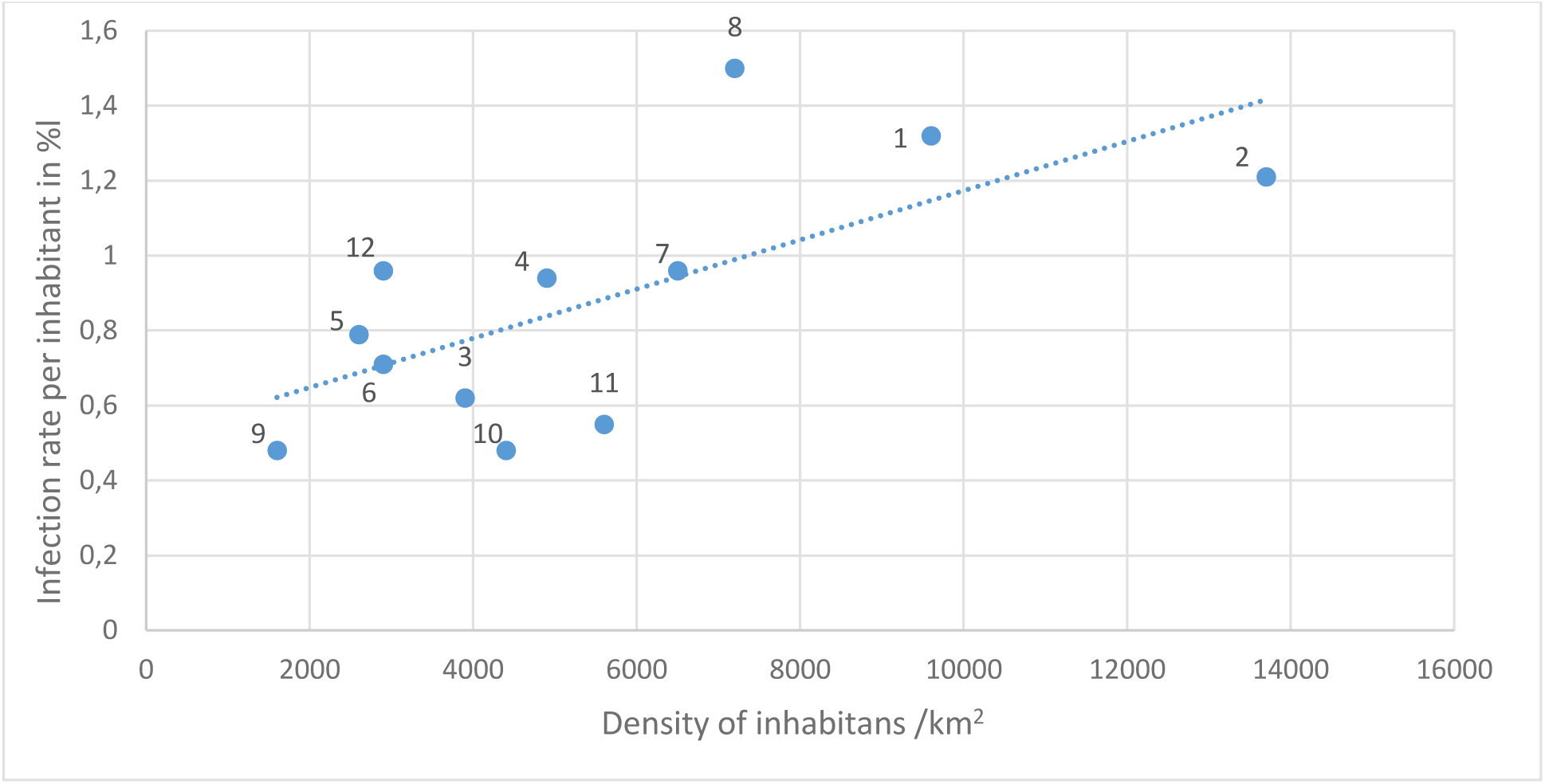
The dependence of infection rate on density of inhabitants.

Analysis of this graph shows that this statistical relationship is indeed found: as the population density increases, the relative number of infected people increases.

However, for regions with the highest number of infected people, the population density is higher than for other districts in Berlin. But this relationship is much lower than that shown in Fig. 6 (correlation coefficient r = 0.67).

Fig. 4, Fig. 5 and Fig. 6 show the districts of Berlin in numbers respectively:

1- Mitte; 2- Friedrichshain–Kreuzberg; 3- Pankow; 4- Charlottenburg–Wilmersdorf; 5- Spandau; 6 - Steglitz–Zehlendorf; 7- Tempelhof–Schöneberg; 8- Neukölln; 9- Treptow–Köpenick; 10- Marzahn–Hellersdorf ; 11- Lichtenberg ; 11- Reinickendorf.

Another important factor that affects the rate of spread of infection is the age of infected persons.

Figure 7 shows the relative number of people infected with coronavirus according to their age. The maximum percentage of infected people are young people between the ages of 20 and 29 who are the most socially active. The disease rate in this age group reaches 1.5%. School-age children and people in the 30 to 50 age group are slightly weaker, slightly more than 1%. For these major age groups, restrictions on social contacts must first be introduced in order to slow down the spread of the epidemic.

**Figure 7:**
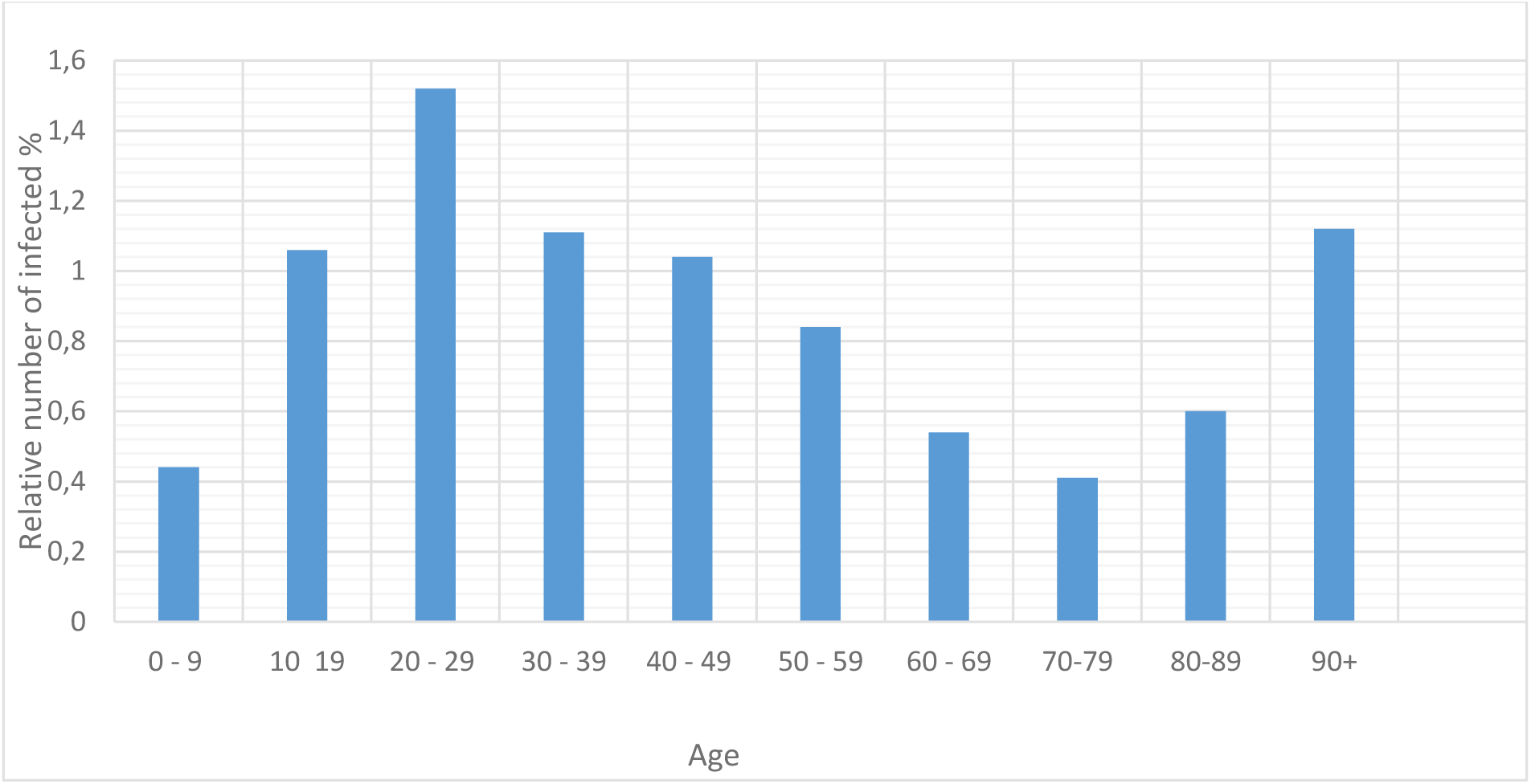
Relative number of infected people according to their age.

This data already makes it possible to develop more effective measures to limit the growth of the epidemic, targeting primarily those groups of the population that are more heavily infected.

In addition to the socio-demographic characteristics of the city’s population, the spread of the epidemic may also be influenced by the psychological state of people who determine their behavior under the epidemic, i.e. their readiness to comply with hygienic or social rules of distance.

Identifying psychological factors that may encourage engagement in preventative health behaviours is crucial. The behavioural immune system (BIS) represents a set of psychological processes thought to promote health by encouraging disease avoidance behaviours.

A set of psychological factors that may be particularly relevant for understanding responses to COVID-19 comes from theory on the behavioural immune system (BIS) [6]. Those higher in BIS reactivity should be more sensitive to pathogen threats and more avoidant of potential contamination. As such, individuals higher in disgust sensitivity and germ aversion should be more concerned with the COVID-19 pandemic and more likely to engage in preventative health behaviours to avoid the disease.

Neutralizing pathogens requires more than detection; it also requires avoidance. The emotion disgust (and, specifically, pathogen disgust) serves this function [7]

An important objective indicator of the level of disgust, as well as other major emotions - fear, anxiety or stress - can be the analysis of changes in heart rate. This physiological measure can provide information about how quickly and well the body adapts to external influences. Heart rate variability (HRV) is the variance in the intervals (differences in duration) between heartbeats over time. HRV analysis is the ability to assess general heart health and the condition of the autonomic nervous system (ANS), which is responsible for regulating heart activity. The ANS is divided into two parts: The sympathetic and parasympathetic systems. The sympathetic system ensures maximum performance. The parasympathetic system, on the other hand, is used for recovery, relaxation and the restoration of strength after heavy exertion, which is activated by the sympathetic system.

There should be a balance between these two areas to ensure optimal health. Through the sympathetic system, goals are achieved and performance is achieved, but this can only continue if the parasympathetic system recovers the body from this work in time.

The spikes of the heartbeats (the upward rash on an ECG) are called R-spikes and the intervals between the R-spikes are called RR intervals.

The LF (“low frequency”: 0.04- 0.15 Hz) is described by the longer-term changes in successive RR intervals. Both the sympathetic and parasympathetic systems have an influence on the LF, but the sympathetic system has a greater effect. HF (“high frequency”: 0.15- 0.4 HZ) represents the short-term changes in successive RR intervals and is a sound measure of parasympathetic activation. The LF/HF ratio describes the quotient of LF and HF. This value is often used to measure the balance in the ANS between sympathetic and parasympathetic activation.

Experimental and theoretical studies were carried out at the workshop of D. Below at the University of Potsdam to study the relationship between disgust response and HRV characteristics [8].

The aim of this study was to see if disgust could be detected using heart rate variability and used for diagnostic purposes in the future. 30 students from the University of Potsdam took part in this study. Images and objects were to be evaluated by the test persons according to their subjective level of disgust, contamination, threat and anxiety on a self-generated scale. The heart rate variability was measured during the presentation of the objects and images and a 5 minute pre-measurement (baseline).

Images and objects were to be rated by the test persons according to their subjective disgust, contamination, threat and fear content on a self-created scale.

The coefficient of resistance B is used as a characteristic value reflecting the intensity of changes in the physiological dimensions of the heart. This coefficient is equal to the relationship between a physiological measurement of the heart at rest and the same physiological measurement during psychological stress, in this case disgust. This resistance coefficient B was previously shown as a quotient in the calculations. To simplify matters, from now on only the resistance coefficient B will be used, as only the ratio of HF during disgust induction and HF during the resting phase is calculated. A formula has been drawn up for this:

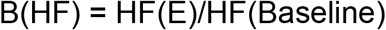

Figure 8 shows the results of the dependence of the resistance coefficient B (HF) for the presentation phase of the images on the ratio during baseline measurement. The correlation coefficient between B for the presentation phase of the images and the ratio at rest is significant (r = .623, p < .01).

**Fig. 8.**
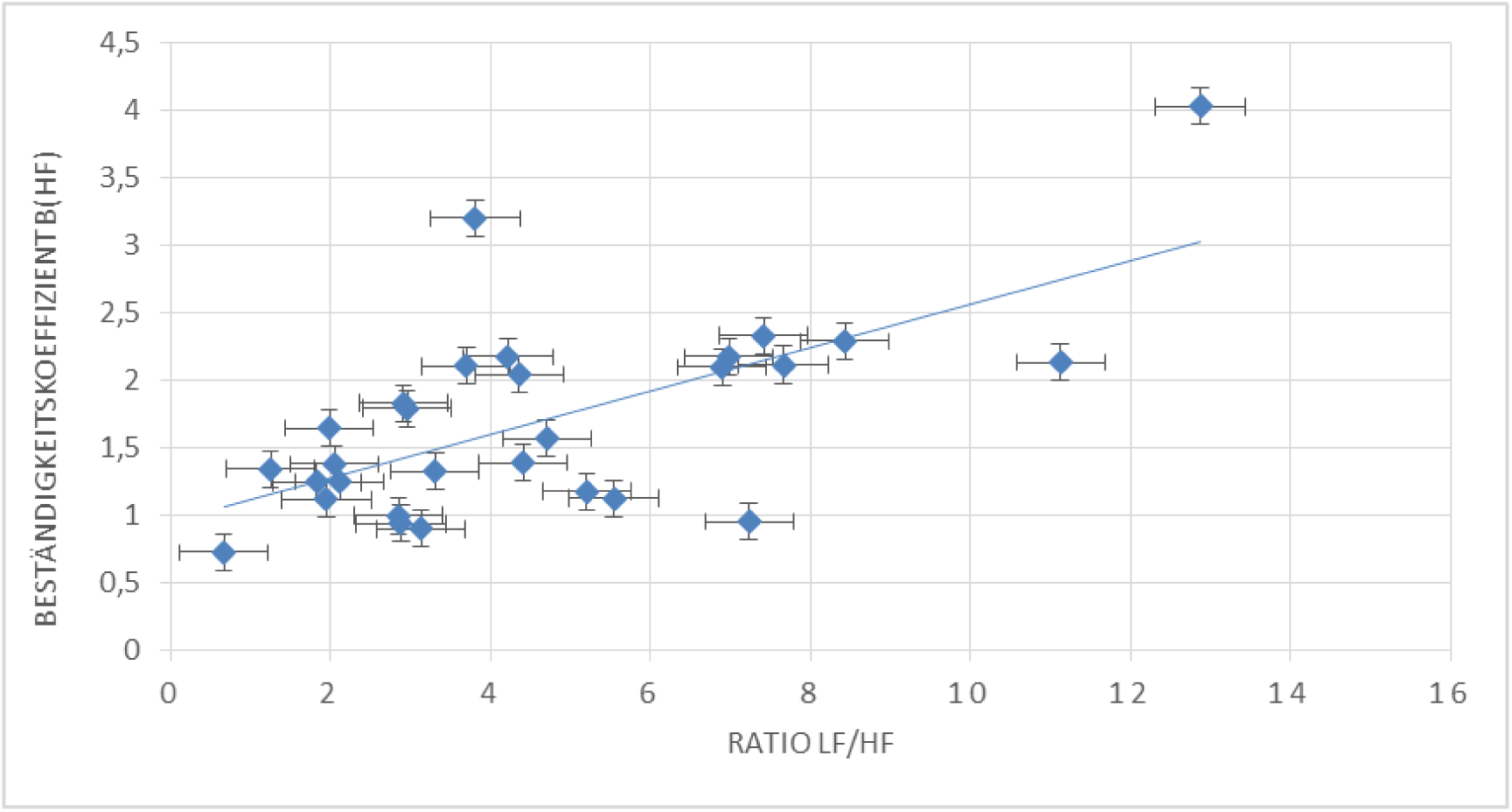
Dependence of B coefficient on LF/HF ratio

A similar schedule has also been obtained in experiments with objects. Between these two parameters, ratio and resistance coefficient B (HF), a relationship is shown in the form of a formula. This formula is:

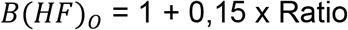

HF(E) is the HF during the presentation phase while the HF(Baseline) is the HF during the baseline measurement.

By introducing the resistance coefficient B (HF), the physiological reaction to a feeling of disgust or stress can thus be predicted. This requires measuring the ratio at rest, as was done in this study during baseline measurement. Inserting this value in the above formulae in relation to the type of presentation that would help to trigger the feeling of disgust or stress, gives the expected value for the resistance coefficient B (HF) for the induction method in question. If, furthermore, the HF values in the resting phase are known, the expected HF value during a disgust or stress induction phase can be calculated using the following formula,

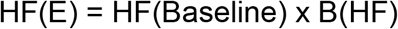

To further increase the usefulness of these derivatives, a more detailed analysis of the resistance coefficient B (HF) would have to be carried out, depending on the intensity of the disgust or stress caused, age, sex and state of health (taking medication or the presence of a disease). Furthermore, the link between the resistance coefficient B (HF) and the stress index must be strengthened in order to predict the change in disgust or stress even more accurately, as the stress index reflects the balance in the ANS even more accurately. Lastly, research must be carried out into differences and similarities in the response of stress and disgust in relation to physiological measures, so that the derived formulas and procedures can be used definitively in stress and disgust.

An attempt to link the results of the HRV analysis with subjective data from the survey of the participants in the experiment showed that there is no correlation between them, which is the advantage of the B coefficient, which objectively reflects the degree of impact of disgust on the psyche of the participants, and the subjective assessment of such impact based on data from special questionnaires.

The use of parameter B as the main characteristic of the activity of the nervous system’s response to external influences and the relationship of this parameter to the LF/HF ratio at rest can be successfully used in the analysis of other types of psychological disorders, such as depression, increased stress levels, etc., in particular, in conditions of additional influences related to both the need for quarantine and the risk of disease. In general, it can be predicted that with the increase in the LF/HF ratio, the intensity of exposure related to COVID-19 increases accordingly.

At the same time, as this relationship grows, the level of psychological reaction to external influences increases, i.e. the intensity of both anxiety and feelings of disgust increases. And these emotions, as already mentioned, form the basis of the BIS behavioural profile that can influence the spread of the COVID-19 epidemic.

Special research must be carried out to confirm that the intensity of the epidemic may depend on the psychological state of the population. However, it is safe to assume that the psychological state of the population should be taken into account when introducing restrictive measures to reduce the epidemic. Otherwise, the effectiveness of such measures may not be high enough. In the same way that so-called economic psychology has recently begun to develop, the new conditions associated with the global spread of epidemics require detailed study of previously unknown problems of epidemiological psychology.

In general, when looking at the nature of the growth of diseases, it must be recognised that restrictive measures do not lead to an effective result, as they are implemented without analysing and taking into account the causal links that determine the extent to which individual factors influence the intensity of the spread of the epidemic. For example, for Berlin, instead of strengthening health controls, the city administration has ordered the complete closure of restaurants or gyms, for example, for playing tennis. At the same time, the most likely places of transmission, such as schools or public transport, continue to operate as usual. Moreover, instead of reducing contact levels as much as possible by maximising the number of trains and buses during the epidemic, a number of routes were cancelled over several weeks. The lack of scientific analysis of the most active sources and routes of infection does not make it possible to carry out optimal management of the city during the epidemic in terms of protecting public health and maintaining the economic standard of living.

In addition to the conclusions drawn in the first part of this paper [1], the following conclusions can be added:

1. The proposed model adequately describes the development of the coronavirus epidemic with insufficient adherence to quarantine and social distancing. It shows that with an epidemic growth rate of around 1,000 people/day, unless additional quarantine measures are taken, the total number of infections can be expected to approach 100,000 within approximately six months.
2. The further development of the model should thus go in the direction of identifying causal links between the intensity of the epidemic and the main factors affecting this process. Some of these factors are related to the characteristics of the population’s behaviour and the infrastructure of cities.
3. The increase in the incidence in areas with a large percentage of the population rooted in Islamic countries is more than 3 times higher than in the rest of Berlin.
4. An additional factor affecting the intensity of the epidemic is population density. As the population density increases, the incidence increases.
5. The age composition of the population also affects the speed at which the epidemic spreads. The maximum relative infection of the virus is observed in young people between the ages of 20 and 30.
6. One of the factors influencing the intensity of the epidemic’s growth may be the psychological resilience of the population associated with the BIS concept of behavioural immunity. Preliminary results of the study show that the level of intensity of the main emotions that determine the BIS (primarily the level of disgust) can be linked to the characteristics of the heart rate. A methodology has been proposed to predict the possible reaction of individuals to the epidemic based on an analysis of their heart rate.
7. The effectiveness of anti-epidemiological measures can be improved by quarantine measures primarily for those groups of the population most heavily infected, with the possibility of introducing different restrictive measures for each district of even one city.
8. The lack of scientific analysis of the most active sources and routes of infection does not make it possible to carry out optimal management of the city during the epidemic in terms of protecting public health and maintaining the economic standard of living.

## Data Availability

[1] Below, D., & Mairanowski, F. (2020). Prediction of the coronavirus epidemic prevalence in quarantine conditions based on an approximate calculation model. medRxiv. [2] Hodcroft, E. B., Zuber, M., Nadeau, S., Comas, I., Candelas, F. G., Stadler, T., & Neher, R. A. (2020). Emergence and spread of a SARS-CoV-2 variant through Europe in the summer of 2020. medRxiv. [3] Der Regierende Buergermeister von Berlin, Senatskanzlei Informationen zum Coronavirus (Covid-19). https://www.berlin.de/corona/fallstatistik. [4] Amt fuer Statistik Berlin-Brandenburg (2020). Statistischer Bericht A I 16, hj 2/ 19 Einwohnerinnen und Einwohner im Land Berlin am 31. Dezember 2019. Potsdam. [5] Unternehmenservice Berliner Bezirke. (05.11.2020). https://www.businesslocationcenter.de/wirtschaftsstandort/berlin-im-ueberblick/berliner-bezirke/. [6] Shook, N. J., Sevi, B., Lee, J., Oosterhoff, B., & Fitzgerald, H. N. (2020). Disease avoidance in the time of COVID-19: The behavioral immune system is associated with concern and preventative health behaviors. PloS one, 15(8), e0238015. [7] Tybur, J. M. (2020). How Coronavirus Bypasses Our Behavioral Immune System (And What We Can Do About It). https://thisviewoflife.com/how-coronavirus-bypasses-our-behavioral-immune-system-and-what-we-can-do-about-it/. Published: 23.03.2020.

